# Machine learning to predict 5-year survival among pediatric Acute Myeloid Leukemia patients and development of OSPAM-C online survival prediction tool

**DOI:** 10.1101/2020.04.16.20068221

**Authors:** Ashis Kumar Das, Shiba Mishra, Devi Kalyan Mishra, Saji Saraswathy Gopalan

## Abstract

**Background:** Acute myeloid leukemia (AML) accounts for a fifth of childhood leukemia. Although survival rates for AML have greatly improved over the past few decades, they vary depending on demographic and AML type factors.

**Objectives:** To predict the five-year survival among pediatric AML patients using machine learning algorithms and deploy the best performing algorithm as an online survival prediction tool.

**Materials and methods:** Pediatric patients (0 to 14 years) with a microscopically confirmed AML were extracted from the Surveillance Epidemiology and End Results (SEER) database (2000-2011) and randomly split into training and test datasets (80/20 ratio). Four machine learning algorithms (logistic regression, support vector machine, gradient boosting, and K nearest neighbor) were trained on features to predict five-year survival. Performances of the algorithms were compared, and the best performing algorithm was deployed as an online prediction tool.

**Results:** A total of 1,477 patients met our inclusion criteria. The gradient boosting algorithm was the best performer in terms of discrimination and predictive ability. It was deployed as the online survival prediction tool named OSPAM-C (https://ashis-das.shinyapps.io/ospam/).

**Conclusions:** Our study provides a framework for the development and deployment of an online survival prediction tool for pediatric patients with AML. While external validation is needed, our survival prediction tool presents an opportunity to reach informed clinical decision-making for AML patients.

## 1. Introduction

Acute myeloid leukemia (AML) is a heterogenous hematological cancer with expansion of abnormally differentiated myeloid hematopoietic progenitor cells and it accounts for a fifth of childhood leukemia [1,2]. The overall survival of children due to AML has improved in the recent decades due to advancements in therapy and it is currently around 70% [3–5]. However, survival rates vary depending on demographic and AML type factors [6–9]. Therefore, it is essential to understand the prognostic factors for AML outcomes for effective planning of treatment and rehabilitation modalities. While there have been few studies translating the prognostic factors to predictive models on AML, they have focused on adult patients and none have used machine learning specifically for predicting pediatric patient survival [10,11].

Machine learning consists of a group of artificial intelligence techniques, where the algorithms learn the patterns in the data without being explicitly programmed to carry out specific applications. Learning from a set of data (training data), machine learning algorithms apply a predictive model to unseen data (test data) [12]. Utilizing the already available data from hospitals and medical databases, machine learning has the potential to diagnose health conditions, predict appropriate treatment methods and patient survival to improve overall quality of life. There have been several applications of machine learning in healthcare, such as predicting diseases, health events and drug response, survival prediction, clustering of patients based on risk classification, analyzing genetics data and medical imaging [13–17]. In the field of cancer research, a few studies have utilized machine learning for predicting cancer survival from hospital records and registries [18–23]. The Surveillance Epidemiology and End Result (SEER) database is the largest publicly available source of cancer statistics in the United States and it includes approximately 28% of the population [24]. Though several studies have applied machine learning on predicting patient survival on various cancers from SEER database, none have applied it on AML for pediatric patients [20–22].

Our study had two objectives, (1) predict the five-year survival among pediatric (0 to 14 years) AML patients using machine learning algorithms, and (2) deploy the best performing algorithm as a web application for future validation and clinical use.

## 2. Material and methods

### 2.1. Patients

Patients for this study were selected from the Surveillance Epidemiology and End Result (SEER) database (1975-2016) [25]. The standard for case completeness for the SEER database is 98% and all patients were followed up for 10 years after routine treatment until death or loss to follow-up [26]. The database includes patient details from 1975 through 2016 and reports their demographic background, cancer characteristics, and survival. The available variables on AML were age, sex, race, marital status, AML histologic subtype, AML grade, SEER registry details (name, state and county), year of diagnosis, and survival in months.

Our inclusion criteria for this study were microscopically confirmed AML for patients aged 14 or younger. We excluded patients without microscopically confirmed AML, with unknown survival time and those with their years of diagnosis before 2000. So as to have adequate follow up period after the diagnosis, we considered the patients diagnosed between 2000 and 2011 as our sample. A total of 76,382 AML patients were diagnosed with AML between 1975 and 2016 across all age groups. After excluding patients that did not meet our inclusion criteria, 1,477 pediatric AML patients were included in our study.

### 2.2. Outcome variable

Our outcome variable was survival of five years or more among AML patients. In the SEER database, survival is a continuous variable with units in months. So, we created a binary variable where any patient with a survival of 60 months or more was coded “yes”, or otherwise “no”.

### 2.3 Predictors

We considered individual patient level demographic and disease variables as predictors. Demographic predictors were sex, age (years at diagnosis), and race. There were six races – “Hispanic”, “non-Hispanic American Indian/Alaska native”, “non-Hispanic Asian or Pacific Islander”, “non-Hispanic black”, “non-Hispanic white” and “non-Hispanic unknown”.

Disease variables that were available in the database were AML sub-type and grade. In our sample, there were 14 AML subtypes available according to the 3rd edition of the International Classification of Diseases for Oncology (ICD-O-3) [27]. The AML subtypes were the following: 9840/3 – acute erythroid leukemia; 9861/3 – AML, NOS; 9866/3 – acute promyelocytic leukemia (AML with t (15;17) (q22; q12)) PML/RARA; 9867/3 – acute myelomonocytic leukemia; 9871/3 – AML with inv (16)(p13.1q22) or t (16;16) (p13.1;q22), CBFB-MYH11; 9872/3 – AML with minimal differentiation; 9873/3 – AML without maturation; 9874/3 – AML with maturation; 9895/3 – AML with myelodysplasia-related changes; 9896/3 – AML, t (8;21)(q22;q22) RUNX1-RUNX1T1; 9897/3 – AML with t (9;11) (p22;q23), MLLT3-MLL; 9898/3 – AML with Down Syndrome; 9910/3 – acute megakaryoblastic leukemia; and 9920/3 – therapy related myeloid neoplasm. A vast majority of patients (93 percent) had unknown AML grade. Thus, we excluded this variable from our analysis.

### 2.4 Statistical Methods

#### 2.4.1 Descriptive Analysis

We performed descriptive analyses for the predictors stratified by their classes. The correlation was tested among all predictors with Pearson’s correlation coefficient.

#### 2.4.2 Predictive Analysis

We employed machine learning to predict the determinants of five-year survival to AML. We applied four commonly used supervised machine learning algorithms in cancer research – logistic regression, support vector machine, K neighbor classification, and gradient boosting – to understand which algorithm provides higher accuracy of prediction. We ran the best-fitting model for each algorithm to derive the predictions. The best-fit was derived through optimization techniques as described under each algorithm below.

##### 2.4.2.1 Logistic Regression (LR)

Logistic regression is used for classification problems, i.e. binary or categorical output. The algorithm fits the best model to describe the relationship between the output and input (predictor) variables [28]. We used the grid search function to identify the best fit parameters, which were L2 regularization and a penalty strength of 1.

##### 2.4.2.2 Support Vector Machine (SVM)

The data is classified into two classes in support vector machine (SVM) based on the output variable over a hyperplane [23]. The algorithm tries to maximize the distance between the hyperplane and the two closest data points from each class. There are three critical parameters in SVM – kernel (transforms data into a spatial form such as linear, radial, sigmoid, or polynomial), penalty (an error term, also called regularization) and gamma (a measure of model fitting). Using grid search feature for optimization, the best parameters in our model for kernel, penalty and gamma were radial, 1 and 0.1 respectively.

##### 2.4.2.3 K Nearest Neighbors (KNN)

The class of a new observation is decided by the majority class among its neighbors in KNN algorithm [29]. There are three important parameters for KNN – number of nearest neighbors, distance metric and weights. Number of nearest neighbors refers to the number of data points a new observation is assigned to. Distance metric is a measure of the distance between the new observation and the nearest neighbors. There are three possible distance metrics – Euclidean, Manhattan and Minkowski. Weight is a measure to test the contribution of the members in the neighborhood. The members can be weighted equally (uniform weight) or higher weights for nearest members (distance weight). Using grid search feature for optimization, the best parameters in our model were 15 nearest neighbors, Manhattan metric and uniform weights.

##### 2.4.2.4 Gradient Boosting

Gradient boosting is an algorithm that uses a combination of shallow and successive decision trees [30]. Decision trees consist of recursively partitioning (also known as splitting) of the predictors. Each decision tree learns successively and improves on the previous (learning rate). One must define the maximum depth for each decision tree (number of levels up to which splitting continues) and minimum leaf sample to split (minimum number of observations required in a node to be considered for splitting). Eventually, predictions are based on a weighted combination of these trees. We used grid search feature to optimize model parameters. The best fit parameters were – 80 decision trees, maximum depth of three for each tree, minimum leaf samples of seven to split, three maximum features and 0.15 learning rate.

##### 2.4.2.5 Evaluation of the performance of the algorithms

The data was split into training (80 percent) and test segments (20 percent) for all algorithms. First, the algorithms were trained on the training segment and then were validated on the test segment for determining predictions. The data was 10-fold cross-validated with the data split into 80% training and 20% test observations randomly ten times for all algorithms. The average of the cross-validations was taken as the final result. The models were evaluated with accuracy (correct prediction of survived patients as survived and non-survived patients as non-survived), precision (ratio of correctly predicted survived patients to the total predicted survived patients), recall (ratio of correctly predicted survived patients to the all patients), F1 score (weighted average of precision and recall), and area under the receiver operating characteristics curve (AUC) [35]. A receiver operator characteristic (ROC) curve presents a plot of the true positive rate (y-axis) against the false positive rate (x-axis) for each individual algorithm. AUC measures the area under the ROC curve, and it ranges from 0.50 to 1.0 where 0.50 indicates the lowest discriminating score and 1.0 indicates the highest discriminating score.

The statistical analyses were performed using Python programming language Version 3.7 (Python Software Foundation, Wilmington, DE, USA) and the deep neural network was implemented on the TensorFlow platform [36]. The web application was built using the Shiny package for R and deployed with Shiny server (R Foundation for Statistical Computing, Vienna, Austria).

## 3. Results

In this section, we present the profile of patients, performance of the algorithms and our online survival prediction tool.

### 3.1 Patient profile

The demographic profile of the patients is presented in Table 1. The mean age of the patients was 6.1 years with a standard deviation of 5. Slightly above half were males (52.9%). Among various races, non-Hispanic whites were the majority (43.4%) followed by Hispanics (31.8%) and non-Hispanic blacks (13.7%). Out of all AML subtypes, patients with AML not otherwise specified (NOS) were the majority group (39.2%). Closer to 60% of the patients in our sample had a survival of five or more years. The correlation coefficients between the predictors ranged from – 0.14 to 0.02.

**Table 1.**
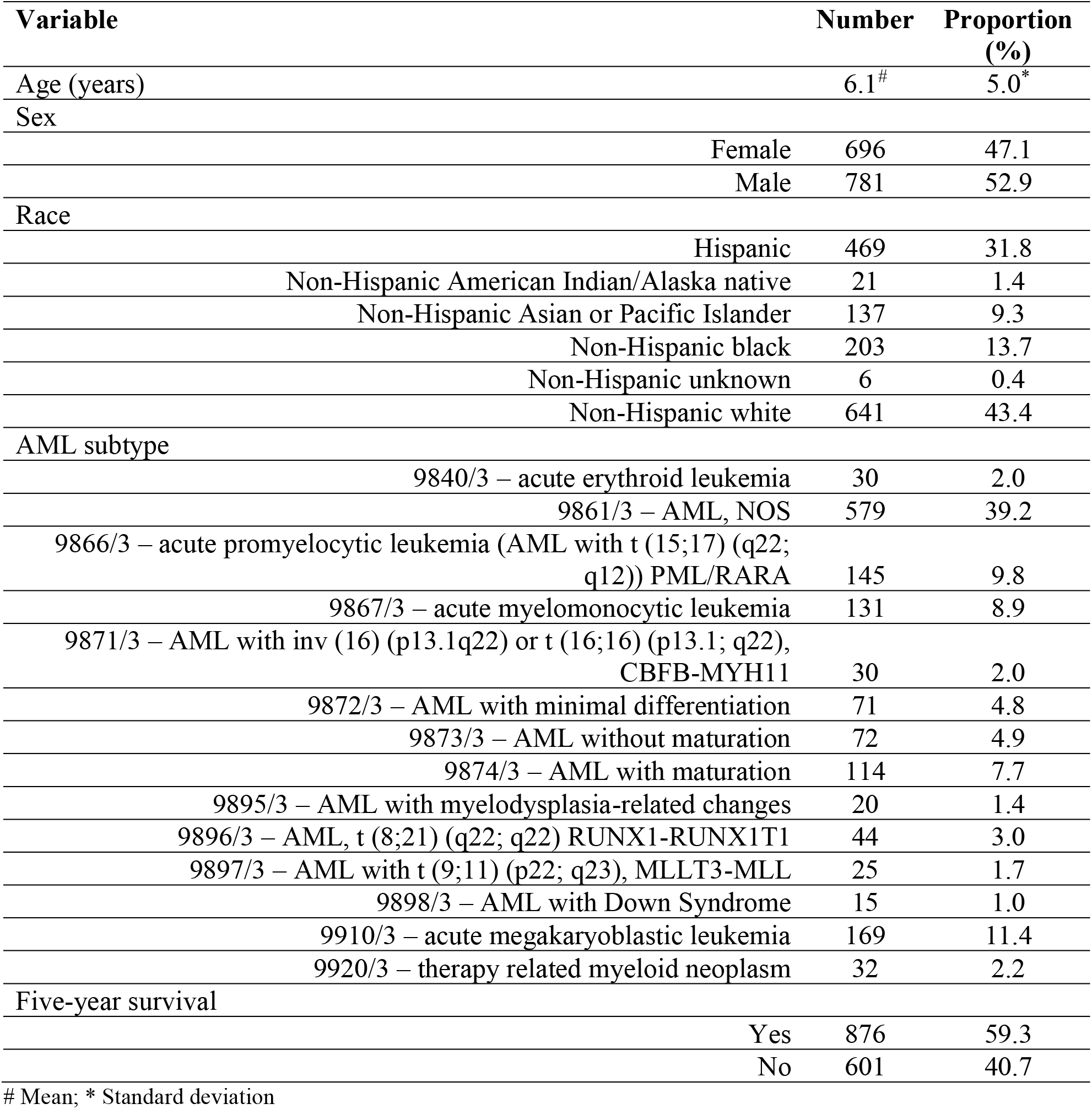
Patient profile

| Variable | Number | Proportion (%) |
| --- | --- | --- |
| Age (years) | 6.1<sup>#</sup> | 5.0<sup>*</sup> |
| Sex |  |  |
| Female | 696 | 47.1 |
| Male | 781 | 52.9 |
| Race |  |  |
| Hispanic | 469 | 31.8 |
| Non-Hispanic American Indian/Alaska native | 21 | 1.4 |
| Non-Hispanic Asian or Pacific Islander | 137 | 9.3 |
| Non-Hispanic black | 203 | 13.7 |
| Non-Hispanic unknown | 6 | 0.4 |
| Non-Hispanic white | 641 | 43.4 |
| AML subtype |  |  |
| 9840/3 – acute erythroid leukemia | 30 | 2.0 |
| 9861/3 – AML, NOS | 579 | 39.2 |
| 9866/3 – acute promyelocytic leukemia (AML with t (15;17) (q22; q12)) PML/RARA | 145 | 9.8 |
| 9867/3 – acute myelomonocytic leukemia | 131 | 8.9 |
| 9871/3 – AML with inv (16) (p13.1q22) or t (16;16) (p13.1; q22), CBFB-MYH11 | 30 | 2.0 |
| 9872/3 – AML with minimal differentiation | 71 | 4.8 |
| 9873/3 – AML without maturation | 72 | 4.9 |
| 9874/3 – AML with maturation | 114 | 7.7 |
| 9895/3 – AML with myelodysplasia-related changes | 20 | 1.4 |
| 9896/3 – AML, t (8;21) (q22; q22) RUNX1-RUNX1T1 | 44 | 3.0 |
| 9897/3 – AML with t (9;11) (p22; q23), MLLT3-MLL | 25 | 1.7 |
| 9898/3 – AML with Down Syndrome | 15 | 1.0 |
| 9910/3 – acute megakaryoblastic leukemia | 169 | 11.4 |
| 9920/3 – therapy related myeloid neoplasm | 32 | 2.2 |
| Five-year survival |  |  |
| Yes | 876 | 59.3 |
| No | 601 | 40.7 |

### 3.2 Performance of the algorithms

The performance metrics of the algorithms (logistic regression, support vector machine, K nearest neighbor, and gradient boosting) are shown in table 2. The accuracy of gradient boosting was the highest (0.681) followed by KNN (0.635), SVM (0.618), and logistic regression (0.588). F1-score (harmonic mean of precision and recall) was the highest for the gradient boosting (0.692), followed by SVM (0.672), logistic regression (0.664), and KNN (0.663). Area under receiver operating characteristic curve (AUC) ranged from 0.561 to 0.726 with the highest score for the gradient boosting algorithm. Considering all the performance metrics, gradient boosting was the best performer.

**Table 2.** Performance metrics of the algorithms

| **Metrics** | **Logistic regression** | **Support vector machine** | **K nearest neighbor** | **Gradient boosting** |
| --- | --- | --- | --- | --- |
| Accuracy (SD*) | 0.588 (0.077) | 0.618 (0.047) | 0.635 (0.037) | 0.681 (0.047) |
| Precision | 0.721 | 0.713 | 0.688 | 0.692 |
| Recall | 0.615 | 0.636 | 0.639 | 0.692 |
| F1-score | 0.664 | 0.672 | 0.663 | 0.692 |
| AUC | 0.635 | 0.561 | 0.665 | 0.726 |

\* Standard deviation of 10-fold cross-validated accuracy

### 3.3 Online survival prediction tool – OSPAM-C

The best performing model, gradient boosting was deployed as the online survival prediction tool named as “**O**nline **S**urvival **P**rediction tool for **A**cute **M**yeloid Leukemia in **c**hildren” – “OSPAM-C” (https://ashis-das.shinyapps.io/ospam/). As shown in figure 1, the user interface has four boxes to select input features as drop-down menus. The features are age (fourteen options – 0 through 14 years), sex (two options – male and female), race (six options – Hispanic, non-Hispanic American Indian/Alaska native, non-Hispanic Asian or Pacific Islander, non-Hispanic Black, non-Hispanic white and unknown) and AML sub-type (seventeen options according to the 3rd edition of the ICD-O-3 and WHO 2008 definitions). A user has to select one option each from the feature boxes and click the submit button to estimate the five-year survival probability in percentages. For instance, the tool gives a five-year survival prediction of 57.4 *%* for a 12-year old female Hispanic patient suffering from AML with maturation (9874/3).

**Figure 1.**
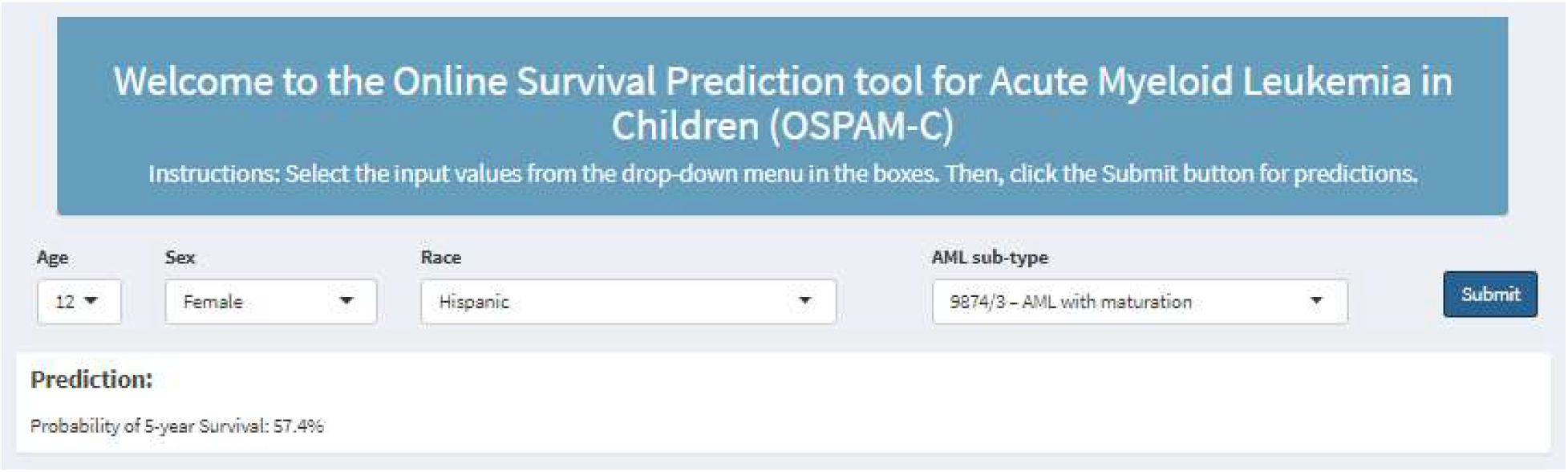
OSPAM-C online survival prediction tool for pediatric AML patients

## Discussion

In this study, we utilized machine learning algorithms to predict five-year survival among pediatric AML patients. Among all our algorithms, gradient boosting performed the best and was deployed as an online survival prediction tool for pediatric AML named OSPAM-C.

Acute myeloid leukemia is one of the most common malignancies among children. While the overall survival has improved for children in recent times, it still has one of the worst survival probabilities among the leading pediatric cancers. AML is also a heterogenous condition with several biological, clinical and genetic factors influencing treatment response and prognosis [37]. While few have explored the predictors of AML survival among children applying conventional analytic methods on SEER database, none have applied machine learning yet [7,38,39].

There are a few predictive web applications to estimate survival for other cancers from SEER database such as chondrosarcoma, spinal chordoma, and glioblastoma [21,40,41]. However, we believe this is the first web-based survival prediction model for pediatric AML patients. Using SEER database, Thio et al. and Karhade et al. applied machine learning algorithms respectively to 1,554 chondrosarcoma and 265 spinal chordoma patients to predict five-year survival [21,40]. They utilized decision tree, support vector machine, Bayes point machine and neural networks. Among their algorithms in both studies, Bayes point machine was the best performer that was deployed for the web application. Similarly, Senders et al applied 15 machine learning and statistical algorithms – accelerated failure time (AFT), bagged decision trees, boosted decision trees, boosted decision trees survival, Cox proportional hazards regression (CPHR), extreme boosted decision trees, k-nearest neighbors, generalized linear models, lasso and elastic-net regularized generalized linear models, multilayer perceptron, naïve Bayes, random forests, random forest survival, recursive partitioning, and support vector machines [41]. The AFT algorithm was deployed as the online prediction tool. The C-statistics (AUC) were 0.868, 0.8 and 0.7 respectively for chondrosarcoma, spinal chordoma, and glioblastoma predictions with their best performing models, whereas it was 0.726 in our best performing model.

Our study has several potential limitations. First, as we used SEER data, there were certain missing clinical features such as treatment type, response to initial therapy, stage and extent of disease. Moreover, due to unavailability of meaningful responses, we had to drop the grade of AML. Second, the database does not collect information on key socio-demographic features such as geographic location, household education and economic status. Third, there was no information in the database on molecular biology, genomics, proteomics, or metabolomics factors. All these additional clinical and socio-demographic factors are known to influence survival in AML patients. Inclusion of these additional features may improve the accuracy and reliability of the model.

Our survival prediction tool is the first of its kind for pediatric AML. Although we used data from the largest cancer database in the US, the tool is yet to be validated. Therefore, we advise caution for clinicians and patients who intend to use this tool as a predictive guide for ascertaining survival for pediatric AML patients. Clinical experts must balance the predictions from this tool against their clinical experience, genomics and other relevant clinical information. We hope this tool will further be validated and possibly reoptimized using heterogenous data from various cohorts in multiple practice settings. While external validation is needed, our survival prediction tool presents an opportunity to inform clinical decision-making for AML patients.

## Data Availability

SEER Data https://seer.cancer.gov/

## Authors’ contributions

Conceived and designed this study: Ashis Kumar Das, Shiba Mishra, Devi Kalyan Mishra, Saji Saraswathy Gopalan

Analyzed and explained the data: Ashis Kumar Das, Shiba Mishra, Devi Kalyan Mishra, Saji Saraswathy Gopalan

All authors contributed to the writing and approved the final manuscript.

## Acknowledgements

We are grateful to the contributors of the Surveillance, Epidemiology, and End Results Program as well as to the National Cancer Institute for making this data publicly available.

## References

[1] A.S. Gamis, T.A. Alonzo, J.P. Perentesis, S. Meshinchi, Children’s Oncology Group’s 2013 blueprint for research: Acute myeloid leukemia, Pediatr. Blood Cancer. (2013). https://doi.org/10.1002/pbc.24432.

[2] B. Deschler, M. Lübbert, Acute myeloid leukemia: Epidemiology and etiology, Cancer. (2006). https://doi.org/10.1002/cncr.22233.

[3] J.R. Davis, D.J. Benjamin, B.A. Jonas, New and emerging therapies for acute myeloid leukaemia, J. Investig. Med. (2018). https://doi.org/10.1136/jim-2018-000807.

[4] S.K. Tasian, J.A. Pollard, R. Aplenc, Molecular therapeutic approaches for pediatric acute myeloid leukemia, Front. Oncol. (2014). https://doi.org/10.3389/fonc.2014.00055.

[5] M. Rasche, M. Zimmermann, L. Borschel, J.P. Bourquin, M. Dworzak, T. Klingebiel, T. Lehrnbecher, U. Creutzig, J.H. Klusmann, D. Reinhardt, Successes and challenges in the treatment of pediatric acute myeloid leukemia: a retrospective analysis of the AML-BFM trials from 1987 to 2012, Leukemia. (2018). https://doi.org/10.1038/s41375-018-0071-7.

[6] U.M. Borate, S. Mineishi, L.J. Costa, Nonbiological factors affecting survival in younger patients with acute myeloid leukemia, Cancer. (2015). https://doi.org/10.1002/cncr.29436.

[7] M.J. Hossain, L. Xie, Sex disparity in childhood and young adult acute myeloid leukemia (AML) survival: Evidence from US population data, Cancer Epidemiol. (2015). https://doi.org/10.1016/j.canep.2015.10.020.

[8] F.R. Appelbaum, H. Gundacker, D.R. Head, M.L. Slovak, C.L. Willman, J.E. Godwin, J.E. Anderson, S.H. Petersdorf, Age and acute myeloid leukemia, Blood. (2006). https://doi.org/10.1182/blood-2005-09-3724.

[9] T. Bochtler, M. Granzow, F. Stölzel, C. Kunz, B. Mohr, M. Kartal-Kaess, K. Hinderhofer, C.E. Heilig, M. Kramer, C. Thiede, V. Endris, M. Kirchner, A. Stenzinger, A. Benner, M. Bornhäuser, G. Ehninger, A.D. Ho, A. Jauch, A. Krämer, Marker chromosomes can arise from chromothripsis and predict adverse prognosis in acute myeloid leukemia, Blood. (2017). https://doi.org/10.1182/blood-2016-09-738161.

[10] C. Chen, P. Wang, C. Wang, Prognostic nomogram for adult patients with acute myeloid leukemia: A SEER database analysis, Medicine (Baltimore). (2019). https://doi.org/10.1097/MD.0000000000015804.

[11] J. Wang, Z. Ma, Q. Wang, Q. Guo, J. Huang, W. Yu, H. Wang, J. Huang, Y. Washington Shao, S. Chen, J. Jin, Prognostic utility of six mutated genes for older patients with acute myeloid leukemia, Int. J. Cancer. (2018). https://doi.org/10.1002/ijc.31178.

[12] L. Liu, Y. Ni, N. Zhang, J. “Nick” Pratap, Mining patient-specific and contextual data with machine learning technologies to predict cancellation of children’s surgery, Int. J. Med. Inform. (2019). https://doi.org/10.1016/j.ijmedinf.2019.06.007.

[13] S. Liu, F. Zhang, L. Xie, Y. Wang, Q. Xiang, Z. Yue, Y. Feng, Y. Yang, J. Li, L. Luo, C. Yu, Machine learning approaches for risk assessment of peripherally inserted Central catheter-related vein thrombosis in hospitalized patients with cancer, Int. J. Med. Inform. (2019). https://doi.org/10.1016/j.ijmedinf.2019.06.001.

[14] Y.J. Tseng, C.E. Huang, C.N. Wen, P.Y. Lai, M.H. Wu, Y.C. Sun, H.Y. Wang, J.J. Lu, Predicting breast cancer metastasis by using serum biomarkers and clinicopathological data with machine learning technologies, Int. J. Med. Inform. (2019). https://doi.org/10.1016/j.ijmedinf.2019.05.003.

[15] Y. Ge, Q. Wang, L. Wang, H. Wu, C. Peng, J. Wang, Y. Xu, G. Xiong, Y. Zhang, Y. Yi, Predicting post-stroke pneumonia using deep neural network approaches, Int. J. Med. Inform. (2019). https://doi.org/10.1016/j.ijmedinf.2019.103986.

[16] D. Ravi, C. Wong, F. Deligianni, M. Berthelot, J. Andreu-Perez, B. Lo, G.Z. Yang, Deep Learning for Health Informatics, IEEE J. Biomed. Heal. Informatics. (2017). https://doi.org/10.1109/JBHI.2016.2636665.

[17] T. Herold, V. Jurinovic, A.M.N. Batcha, S.A. Bamopoulos, M. Rothenberg-Thurley, B. Ksienzyk, L. Hartmann, P.A. Greif, J. Phillippou-Massier, S. Krebs, H. Blum, S. Amler, S. Schneider, N. Konstandin, M.C. Sauerland, D. Görlich, W.E. Berdel, B.J. Wörmann, J. Tischer, M. Subklewe, S.K. Bohlander, J. Braess, W. Hiddemann, K.H. Metzeler, U. Mansmann, K. Spiekermann, A 29-gene and cytogenetic score for the prediction of resistance to induction treatment in acute myeloid leukemia, Haematologica. (2018). https://doi.org/10.3324/haematol.2017.178442.

[18] H. Asri, H. Mousannif, H. Al Moatassime, T. Noel, Using Machine Learning Algorithms for Breast Cancer Risk Prediction and Diagnosis, in: Procedia Comput. Sci., 2016. https://doi.org/10.1016/j.procs.2016.04.224.

[19] K. and S.A. Rajesh, Analysis of SEER Dataset for Breast Cancer Diagnosis using C4.5 Classification Algorithm, Int. J. Adv. Res. Comput. Commun. Eng. (2012).

[20] J.A. Bartholomai, H.B. Frieboes, Lung Cancer Survival Prediction via Machine Learning Regression, Classification, and Statistical Techniques, in: 2018 IEEE Int. Symp. Signal Process. Inf. Technol. ISSPIT 2018, 2019. https://doi.org/10.1109/ISSPIT.2018.8642753.

[21] Q.C.B.S. Thio, A. V. Karhade, P.T. Ogink, K.A. Raskin, K. De Amorim Bernstein, S.A.L. Calderon, J.H. Schwab, Can machine-learning techniques be used for 5-year survival prediction of patients with chondrosarcoma?, Clin. Orthop. Relat. Res. (2018). https://doi.org/10.1097/CORR.0000000000000433.

[22] C.M. Lynch, B. Abdollahi, J.D. Fuqua, A.R. de Carlo, J.A. Bartholomai, R.N. Balgemann, V.H. van Berkel, H.B. Frieboes, Prediction of lung cancer patient survival via supervised machine learning classification techniques, Int. J. Med. Inform. (2017). https://doi.org/10.1016/j.ijmedinf.2017.09.013.

[23] R.O. Alabi, M. Elmusrati, I. Sawazaki-Calone, L.P. Kowalski, C. Haglund, R.D. Coletta, A.A. Mäkitie, T. Salo, A. Almangush, I. Leivo, Comparison of supervised machine learning classification techniques in prediction of locoregional recurrences in early oral tongue cancer, Int. J. Med. Inform. (2020). https://doi.org/10.1016/j.ijmedinf.2019.104068.

[24] S.F. Altekruse, G.E. Rosenfeld, D.M. Carrick, E.J. Pressman, S.D. Schully, L.E. Mechanic, K.A. Cronin, B.Y. Hernandez, C.F. Lynch, W. Cozen, M.J. Khoury, L.T. Penberthy, SEER cancer registry biospecimen research: Yesterday and tomorrow, Cancer Epidemiol. Biomarkers Prev. (2014). https://doi.org/10.1158/1055-9965.EPI-14-0490.

[25] Surveillance, Epidemiology, and End Results (SEER) Program (www.seer.cancer.gov) SEER*Stat Database: Incidence – SEER 9 Regs Research Data, Nov 2018 Sub (1975-2016), National Cancer Institute, DCCPS, Surveillance Research Program, released April 2019, bas, (n.d.).

[26] C. Zippin, D. Lum, B.F. Hankey, Completeness of hospital cancer case reporting from the SEER program of the national cancer institute, Cancer. (1995). https://doi.org/10.1002/1097-0142(19951201)76:11<2343::AID-CNCR2820761124>3.0.CO;2-#.

[27] J.W. Vardiman, J. Thiele, D.A. Arber, R.D. Brunning, M.J. Borowitz, A. Porwit, N.L. Harris, M.M. Le Beau, E. Hellström-Lindberg, A. Tefferi, C.D. Bloomfield, The 2008 revision of the World Health Organization (WHO) classification of myeloid neoplasms and acute leukemia: Rationale and important changes, Blood. (2009). https://doi.org/10.1182/blood-2009-03-209262.

[28] F. Jiang, Y. Jiang, H. Zhi, Y. Dong, H. Li, S. Ma, Y. Wang, Q. Dong, H. Shen, Y. Wang, Artificial intelligence in healthcare: Past, present and future, Stroke Vasc. Neurol. (2017). https://doi.org/10.1136/svn-2017-000101.

[29] H. Raeisi Shahraki, S. Pourahmad, N. Zare, K Important Neighbors: A Novel Approach to Binary Classification in High Dimensional Data, Biomed Res. Int. (2017). https://doi.org/10.1155/2017/7560807.

[30] J. Xie, S. Coggeshall, Prediction of transfers to tertiary care and hospital mortality: A gradient boosting decision tree approach, Stat. Anal. Data Min. (2010). https://doi.org/10.1002/sam.10079.

[31] D.P. Kingma, J.L. Ba, Adam: A method for stochastic gradient descent, ICLR Int. Conf. Learn. Represent. (2015).

[32] G. Klambauer, T. Unterthiner, A. Mayr, S. Hochreiter, Self-normalizing neural networks, in: Adv. Neural Inf. Process. Syst., 2017.

[33] N. Srivastava, G. Hinton, A. Krizhevsky, I. Sutskever, R. Salakhutdinov, Dropout: A simple way to prevent neural networks from overfitting, J. Mach. Learn. Res. (2014).

[34] N. V. Chawla, K.W. Bowyer, L.O. Hall, W.P. Kegelmeyer, SMOTE: Synthetic minority over-sampling technique, J. Artif. Intell. Res. (2002). https://doi.org/10.1613/jair.953.

[35] E.W. Steyerberg, A.J. Vickers, N.R. Cook, T. Gerds, M. Gonen, N. Obuchowski, M.J. Pencina, M.W. Kattan, Assessing the Performance of Prediction Models, Epidemiology. (2010). https://doi.org/10.1097/ede.0b013e3181c30fb2.

[36] M. Abadi, P. Barham, J. Chen, Z. Chen, A. Davis, J. Dean, M. Devin, S. Ghemawat, G. Irving, M. Isard, M. Kudlur, J. Levenberg, R. Monga, S. Moore, D.G. Murray, B. Steiner, P. Tucker, V. Vasudevan, P. Warden, M. Wicke, Y. Yu, X. Zheng, TensorFlow: A system for large-scale machine learning, in: Proc. 12th USENIX Symp. Oper. Syst. Des. Implementation, OSDI 2016, 2016.

[37] U.H. Acharya, A.B. Halpern, Q. (Vicky) Wu, J.M. Voutsinas, R.B. Walter, S. Yun, M. Kanaan, E.H. Estey, Impact of region of diagnosis, ethnicity, age, and gender on survival in acute myeloid leukemia (AML), J. Drug Assess. (2018). https://doi.org/10.1080/21556660.2018.1492925.

[38] S.S. Nasir, S. Giri, S. Nunnery, M.G. Martin, Outcome of Adolescents and Young Adults Compared With Pediatric Patients With Acute Myeloid and Promyelocytic Leukemia, Clin. Lymphoma, Myeloma Leuk. (2017). https://doi.org/10.10167j.clml.2016.09.011.

[39] S. Xie, M.J. Hossain, Survival differences in childhood and young adult acute myeloid leukemia: A cross-national study using US and England data, Cancer Epidemiol. (2018). https://doi.org/10.1016/j.canep.2018.03.001.

[40] A. V. Karhade, Q. Thio, P. Ogink, J. Kim, S. Lozano-Calderon, K. Raskin, J.H. Schwab, Development of Machine Learning Algorithms for Prediction of 5-Year Spinal Chordoma Survival, World Neurosurg. (2018). https://doi.org/10.10167j.wneu.2018.07.276.

[41] J.T. Senders, P. Staples, A. Mehrtash, D.J. Cote, M.J.B. Taphoorn, D.A. Reardon, W.B. Gormley, T.R. Smith, M.L. Broekman, O. Arnaout, An Online Calculator for the Prediction of Survival in Glioblastoma Patients Using Classical Statistics and Machine Learning, Clin. Neurosurg. (2020). https://doi.org/10.1093/neuros/nyz403.

